# Social Networks and Cardiovascular Disease Events in the Jackson Heart Study

**DOI:** 10.1101/2023.03.10.23287131

**Authors:** LáShauntá Glover, Janiyah Sutton, Emily O’Brien, Mario Sims

## Abstract

**Background:** Cardiovascular disease (CVD) disproportionately affects African American adults. Greater social networks (SN), or social connectedness, may lower the risk of CVD events.

**Objective:** Determine the association of SN and incident CVD and test mediation by depressive symptoms, hypertension control and diabetes control.

**Methods:** We used the Social Network Index (SNI) at exam 1 (2000-2004) to develop a continuous standardized SN score and binary categories (high vs. low) among participants in the Jackson Heart Study (n=5252, mean age=54.8 years). Surveillance of coronary heart disease (CHD), stroke, and heart failure (HF) events occurred after exam 1 (2005 for HF) until 2016. Using Cox proportional hazards regression, we estimated the association of SN and CVD events by sex and tested the mediation of depressive symptoms, hypertension control and diabetes control. Models adjusted for age, education, health behaviors, and CVD co-morbidities.

**Results:** Among women, the SN score was associated with a lower risk of CHD and HF after full adjustment (HR 0.78, 95% CI 0.68, 0.89 and HR 0.78, 95% CI 0.63, 0.95, respectively), but the association with stroke attenuated after adjustment for co-morbidities (HR 95% CI 0.88 95% CI 0.67, 1.14). SN scores were also associated with CHD in men (HR 0.84, 95% CI 0.70, 0.99) after full adjustment. High vs. low SN was associated with CHD in men and women, but not after adjustment for co-morbidities. There was no evidence of mediation by depressive symptoms, diabetes control, and hypertension control.

**Conclusion:** Higher SN may lower the risk of CVD events, especially in women.

## Introduction

Every year in the United States, over 600,000 deaths are attributed to cardiovascular disease (CVD), which equates to 1 in 4 deaths.^1^ Age-adjusted death rates for heart disease differ by race and ethnicity, where non-Hispanic Black adults are more likely to have a higher rate than non-Hispanic White, Hispanics and Asian or Pacific Islander adults.^1^ Additionally, the burden of CVD among African American adults is estimated at 9.5%, and the burden of risk factors for CVD, such as hypertension and diabetes, are more common among African American adults than other racial/ethnic groups.^2^

Social isolation, or having few or limited social connections with others, is an upstream social factor associated with inflammation, high cholesterol, and a greater risk of a CVD event.^3 4, 5^ While social isolation is related to a greater risk of CVD, social integration, or having social connections, is associated with a lower risk of CVD.^6^ Social networks (SN), or the social connectedness related to the number, density, and characteristics of family/community connections, may moderate and improve disease risk through a variety of mechanisms. According to Havranek et al.^7^, SN may influence health in two ways: 1) through social influence on behaviors, and 2) through resources embedded in SN that are available to its members. Having a larger and positive SN could also moderate disease risk by reducing certain negative health behaviors such as smoking, by directly or indirectly improving immune system responses, by offering socioeconomic benefits, and by providing greater social support to reduce emotional distress.^8^ Though previous reports have found that cigarette smoking and physical activity are in the pathway of SN and CHD,^6^ it is uncertain whether other factors, strongly associated with CVD are in the pathway, such as having better control of risk factors such as hypertension^9^ and diabetes^10^ and low depressive symptoms.^11^ These key risk factors for CVD may be better managed among those who are better connected and may be the reason for connectedness lowering the risk of CVD.

Social connectedness may be more salient in certain racial ethnic groups. For instance, one study found that African American adults were more likely to be involved in religious organizations when compared to other ethnic groups.^12^ Additionally, African American, compared to White, adults tend to have smaller SN, more family members in their SN, and more frequent contact with their family members.^13^ Due to these differences and a greater burden of CVD risk, there is a need to interrogate how SN relate to CVD endpoints among African American adults in order to better understand what specific areas should be targeted to lower CVD burden and mortality rates.

In this study, SN consisted of five dimensions of social connectedness: marital status, close friends, closeness to relatives, visiting close friends and relatives, and group participation. We tested the association of SN with CVD events (i.e., coronary heart disease (CHD), stroke, and heart failure (HF)) and examined the extent to which depressive symptoms and hypertension control and diabetes control mediated the association of SN with CVD events. Our guiding hypothesis was that there will be an inverse association between SN and incident CVD, with differences among men and women. We also hypothesized that management or control of hypertension and diabetes, and having low depressive symptoms will mediate the association among African American men and women in the Jackson Heart Study (JHS).

## Methods

### Participants

The JHS is a prospective and longitudinal cohort study of CVD among African American adults from the tri-county area (Hinds, Madison, and Rankin) of Jackson, MS. Eligible participants were enrolled at baseline (2000-2004) and followed annually and at each exam [(exam 2 (2005-2008) and exam 3 (2009-2013)]). Participants were sampled from the Atherosclerosis Risk in Communities (ARIC) Study (30%), the Mississippi Department of Transportation Driver’s License and Identification List (17%), volunteers (22%), or family members of those who had already agreed to be a part of the study.^14, 15^ Participants (3,371 women, 1,935 men; age 20-95 years old) provided demographic, socioeconomic, psychosocial, health history, and clinical data from in-home interviews, self-administered questionnaires, and in-clinic examinations at each exam visit. Annual follow up data and surveillance of CVD events were conducted by trained research staff.^16^ All JHS participants provided written informed consent and all study protocols conform to the 1975 Declaration of Helsinki guidelines. The JHS was also approved by the Institutional Review Boards of the following participating institutions: The University of Mississippi Medical Center, Jackson State University, and Tougaloo College.

### Social Networks

We defined SN using five measures based on the definition of social connection and ties from Berkman & Syme Social Networks Index (SNI): marital status, number of close friends, number of close relatives, number of friends or relatives visited monthly, and belonging to social groups.^17^ We generated a continuous SN score based on the five items (range: 0-5) and standardized the score using standard deviation units. To observe threshold effects, we also dichotomized the score as high and low (above and below the median, respectively).

### Mediators

We considered greater hypertension control, diabetes control, and low (vs. high) depressive symptoms as possible mediators in the association between SN and CVD events. Among those with hypertension at exam 1, hypertension control was defined as having a measured blood pressure less than 140/90 mmHg among those treated for hypertension at exam 1. Among those with type 2 diabetes at exam 1, diabetes control was defined as having measured HbA1c levels less than 7.0%. We used the Centers for Epidemiological Studies Depression (CES-D) to define depressive symptoms, which counts the number of clinically significant depressive symptoms experienced during a given week (range 0-60). As recommended, participants who scored greater than or equal to 16 were classified as having high depressive symptoms, while those who scored less than 16 had low depressive symptoms.^18, 19^

### Cardiovascular disease Events

Surveillance of CVD events began during baseline examination (except heart failure, which began in 2005). Surveillance data were available through December 31, 2016. The JHS medical record abstraction team contacted participants by phone to identify CVD events (diagnostic tests, hospitalizations, or death), and also utilized related records (discharge lists and death certificates) from hospitals and state offices for verification.^20, 21^

CHD was defined as definite or probable hospitalized myocardial infarction (MI), a related death, an unrecognized MI by electrocardiogram (ECG), or coronary revascularization based on combinations of chest pain symptoms, ECG changes, and cardiac enzyme levels. A fatal CHD event was classified as an underlying cause of death from the death certificate, and other associated hospital information or medical history.

HF events were defined as the occurrence of either inpatient or outpatient diagnoses of unspecified failure of the heart from an ICD-9 diagnosis code of 428.X in any position or 428 on a death certificate. The definition of HF also includes, but is not limited to, radiographic findings that were similar with congestive HF, increased venous pressure >16 mmHg, dilated ventricle/left ventricular function <40% from an ECG or multiple gated acquisition, or autopsy finding of pulmonary edema.

Stroke events were defined as a definite or probable hospitalized stroke from neuroimaging and autopsy based on the classification from the National Survey of Stroke.^22^ The minimum criterion for probable or definite stroke was rapid onset of neurological symptoms lasting more than 24 hours or symptoms that lead to death. Neurological symptoms lasting less than 24 hours or symptoms seen before or during admission to the hospital were not considered a definite or probable stroke. Only stroke events with medical diagnosis were considered definite or probable. A computer algorithm was used to determine all CVD events, as well as physician reviews. Disagreements were adjudicated by another reviewer.

### Covariates

To determine the inclusion of covariates relevant to the association between SN and CVD, we used a directed acyclic graph (data not shown). Demographic variables such as age (years), sex (male/female), education (<High School Diploma, Some College, College degree or more) were included. Additionally, we included two American Heart Association Life’s Simple 7 characteristics (classified as ideal vs. non-ideal): physical activity and smoking.^23^ Alcohol intake was defined by alcohol drinking in the past 12 months at baseline and calories from fat was defined as daily percent fat from the Food Frequency Questionnaire. CVD comorbidities were also considered: hypertension status (derived variable that includes antihypertensive medication use, self-reported physician diagnosis, and measured systolic and diastolic blood pressure), obesity (based on Body Mass Index >30 kg/m^2^), and diabetes prevalence (derived diabetes status based on ADA 2010 which included fasting glucose ≥ 126 mg/dl, or confirmed medication usage from the medication inventory, or self-reported use of anti-diabetic medications within the past two weeks of the examination, or self-reported diabetes diagnosis).

### Statistical Analysis

There were 5,252 participants who had complete responses from the SNI index at exam 1. Of those, missing observations were removed for education (n=11), smoking (n=79), obesity (n=14), calories from fat (n=177), diabetes prevalence (n=63), and hypertension prevalence (n=2). Because we were interested in incident CVD events, those with baseline CVD were also excluded (n=169), reducing the sample size to 4737. Descriptive statistics, specifically differences in select characteristics by sex, were calculated using chi square and ANOVA tests.

Time-to-CVD event was approximated based on years from exam 1 to the adjudicated incident CVD event (CHD, stroke, HF). Those without CVD and those who had deaths unrelated to CVD were censored. Cox proportional hazards regression was used to estimate the association of high vs. low SN (and the standardized SN score) with incident CVD, where hazard ratios (HR 95% CI) estimated the risk of CVD. Models were sequential in the adjustment of covariates, where Model 1 adjusted for age and educational attainment. Model 2 added physical activity, smoking, alcohol, and dietary fat. Model 3 added hypertension, obesity, diabetes, and depressive symptoms. Because model 3 could include potential mediators, we evaluated these variables by holding them constant when testing for mediation. The proportional hazards assumption was tested by examining Kaplan Meier curves and by examining the statistical significance of time-dependent interaction terms.

Modification by sex was determined by evaluating the interaction term (sex*high SN) in Model 3. A p-value <0.05 was considered statistically significant, as well as confidence intervals without a value of 1.00.

We used Causal Mediation techniques as described by Pearl^24^ and Bind et. al^25^ to relax linearity and interaction assumptions found in the Baron and Kenny method in SAS 9.4 (PROC CAUSALMED) to evaluate mediation. Separate models were constructed to test mediation of hypertension control, diabetes control, and low (vs. high) depressive symptoms respectively in the association between high vs. low SN and each type of CVD event. Models 1 and 2 were used to adjust for confounders, while covariates in Model 3 were not included if the mediator was a covariate. For each potential mediator, we reported the total effect, controlled direct effect, natural direct effect, and the natural indirect effect. Percent mediation was applicable when the natural indirect effect was statistically significant. A p-value <0.05 was considered statistically significant, as well as confidence intervals excluding 1.00.

## Results

**Table 1** shows the descriptive characteristics of participants without CVD at baseline, stratified by sex. Women were older than men (54.9 ±20.6 years vs. 53.5 ± 21.0 years, *p*<0.001), and were more likely to report ideal smoking. Men were more likely to report ideal physical activity (24.2% vs. 17.4%) and were more likely to report alcohol use (59.7% vs. 40.1%). Women were more diabetic (20.9%) and obese (60.1%) and had higher hypertension and diabetes control (25.9% and 14.9%, respectively). Women reported greater depressive symptoms than men (mean=11.2 vs. mean= 10.0, respectively). There were no differences in SN score by sex (*p*=0.607).

**Table 1.** Select characteristics of participants by sex (n=4737), Jackson Heart Study (2000-2004)

| Characteristics | Women (N=3053)<br>N (%) | Men (N=1684)<br>N (%) | Chi Square/ANOVA<br>P-value |
| --- | --- | --- | --- |
| Age (Mean ± SD) | 54.9 ± 20.6 | 53.5 ± 21.0 | <0.001 |
| Education |  |  | <0.264 |
| <High School | 548 (18.0) | 330 (19.6) |  |
| HS-Some College | 1229 (40.4) | 687 (40.8) |  |
| College or Better | 1265 (41.6) | 665 (39.6) |  |
| Smoking |  |  | <0.001 |
| Ideal | 2679 (89.0) | 1344 (81.1) |  |
| Non-Ideal | 330 (11.0) | 314 (18.9) |  |
| Physical Activity |  |  | <0.001 |
| Ideal | 531 (17.4) | 407 (24.2) |  |
| Non-Ideal | 2519 (82.6) | 1275 (75.8) |  |
| Alcohol |  |  | <0.001 |
| Yes | 1217 (40.1) | 1001 (59.7) |  |
| No | 1817 (59.9) | 675 (40.3) |  |
| Dietary fat (Mean ± SD) | 35.1 (7.1) | 35.19 (6.5) | 0.701 |
| Social Networks (range 0-5) |  |  | 0.607 |
| Low (<4) | 509 (16.7) | 271 (16.1) |  |
| High (≥4) | 2544 (83.3) | 1413 (83.9) |  |
| Hypertension | 1322 (55.8) | 671 (53.3) | 0.145 |
| Diabetes | 629 (20.9) | 296 (17.7) | 0.009 |
| Obesity | 1831 (60.1) | 692 (41.2) | <0.001 |
| Hypertension Control | 403 (25.9) | 177 (19.5) | <0.001 |
| Diabetes Control | 418 (14.9) | 182(11.7) | 0.003 |
| Depressive Symptoms | 11.2 ± 6.8 | 10.0 ± 5.9 | <0.001 |

Abbreviations SD, standard deviation.

There were 262 CHD events, 213 stroke events, and 331 HF events over a median follow-up of about 8 years (data not shown). **Table 2** presents the unadjusted cumulative incidence of CVD events by sex. Among women, the cumulative incidence was 5.5% for stroke, 6.0% for CHD, and 8.4% for HF. Among men, the cumulative incidence was 5.8% for stroke, 7.9% for CHD, and 8.5% for HF. The cumulative incidence of HF differed by high and low SN among women. The 10-year unadjusted cumulative incidence of HF was 6.6% (95% CI 5.6%, 7.8%) for women with a high SN score, while the cumulative incidence of HF was 10% (95% CI 7.1%, 13.5%) for women with a low SN score (Gray’s test, p=0.0258). The cumulative incidence of HF did not differ by SN score among men (Gray’s test, p=0.2933) (**Figure 1**). Differences in the cumulative incidence of CHD and stroke by social network score and sex were not statistically significant (**Supplemental Figures A & B**).

**Table 2.**
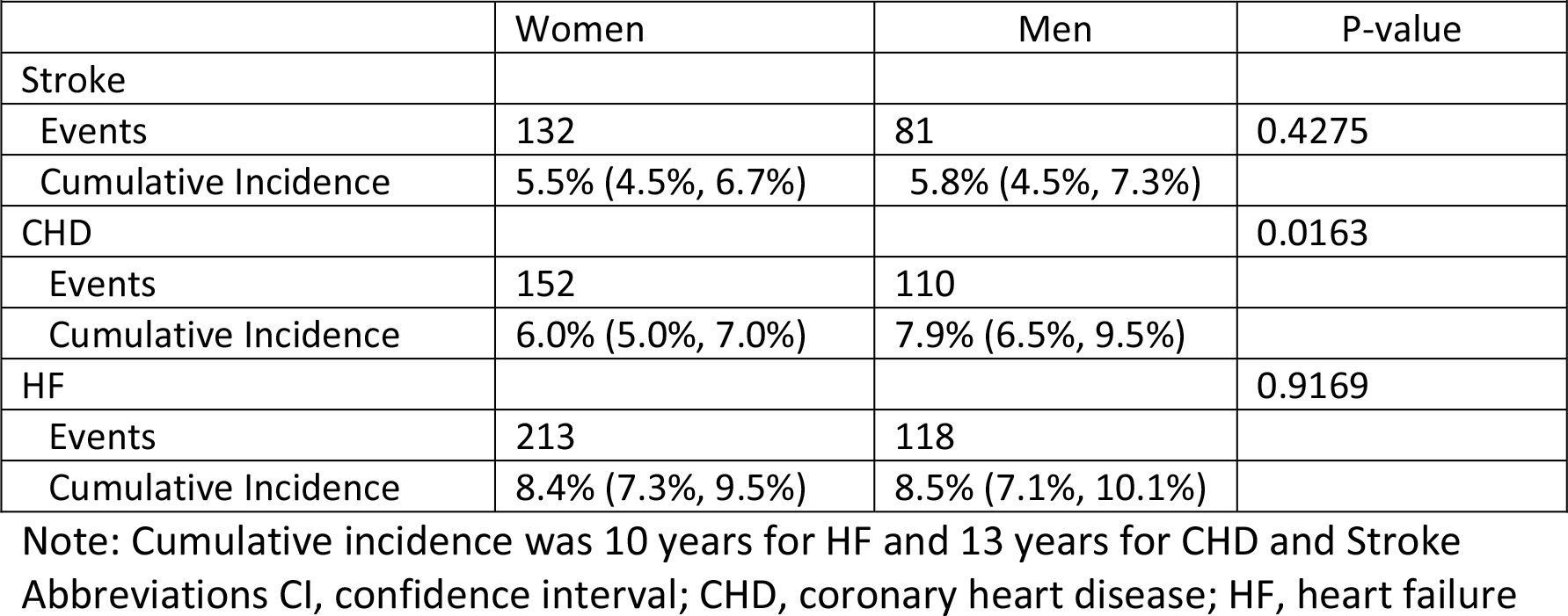
Unadjusted Cumulative Incidence (95% CI) of Cardiovascular Disease Events by Sex

**Figure 1:**
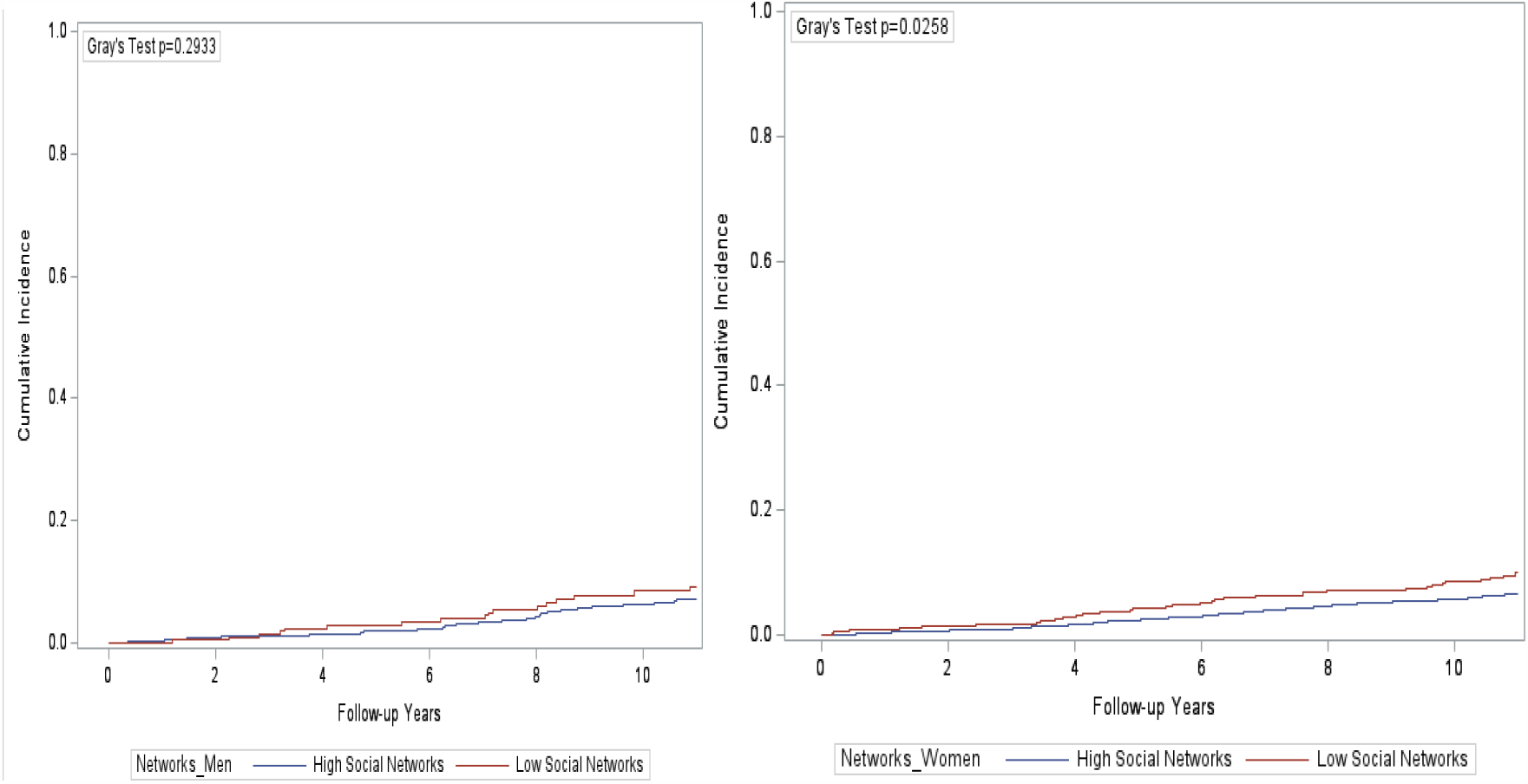
Kaplan-Meier Curve for HF among women (left) and men (right), with high and low social networks.

Table 3 shows the association between SN and CVD events. A 1-SD unit increase in the SN score was associated with a 0.80 (95% CI 0.67, 0.97) hazard of stroke among women in model 2, but the association attenuated in model 3 [HR 0.88 (95% CI 0.67, 1.14)]. Among men and women, a high vs. low social network score was associated with a lower hazard of CHD in model 2 [HR 0.80 (95% CI 0.72, 0.87) and HR 0.84 (95% CI 0.76, 0.93), respectively], and for women and men the association attenuated in model 3 (**Table 3**). Among both men and women, a 1-SD unit increase in SN score was associated with a lower hazard of CHD after full adjustment [women: HR: 0.67 (95% CI 0.54, 0.85); men: HR: 0.69 (95% CI 0.51, 0.93)]. Among women, a high vs. low SN score was associated with a lower hazard for HF in model 2 [HR 0.61 (95% CI 0.43, 0.86)], but the association attenuated after adjustment for covariates in model 3 [HR 0.68 95% CI 0.40, 1.15)]. Additionally, a 1-SD unit increase in the SN score was associated with a lower hazard of HF [HR 0.78 (95% CI 0.63, 0.95)] after full adjustment

**Table 3.** Hazard Ratios (HR, 95% CI) of Cardiovascular Disease Events by Social Networks, stratified by sex: Jackson Study (2000-2016)

| Table 3. Hazard Ratios (HR, 95% CI) of Cardiovascular Disease Events by Social Networks, stratified by sex: Jackson Heart Study (2000-2016) |  |  |  |  |  |  |
| --- | --- | --- | --- | --- | --- | --- |
| STROKE |  |  |  |  |  |  |
|  | Women |  |  | Men |  |  |
|  | Model 1 | Model 2 | Model 3 | Model 1 | Model 2 | Model 3 |
| Social Networks |  |  |  |  |  |  |
| Low (referent) | 1.0 | 1.0 | 1.0 | 1.0 | 1.0 | 1.0 |
| High | 0.71 (0.46, 1.10) | 0.67 (0.43, 1.05) | 0.70 (0.37, 1.34) | 1.23 (0.61, 2.47) | 1.48 (0.70, 3.11) | 1.92 (0.76, 4.84) |
| SD units | **0.81 (0.68, 0.97)** | **0.80 (0.67, 0.97)** | 0.88 (0.67, 1.14) | 0.89 (0.72, 1.09) | 0.95 (0.76, 1.18) | 1.10 (0.74, 1.64) |
| CORONARY HEART DISEASE |  |  |  |  |  |  |
| Social Networks |  |  |  |  |  |  |
| Low (referent) | 1.0 | 1.0 | 1.0 | 1.0 | 1.0 | 1.0 |
| High | **0.64 (0.51, 0.79)** | **0.67 (0.54, 0.85)** | 0.71 (0.50, 1.00) | **0.64 (0.48, 0.85)** | **0.69 (0.51, 0.93)** | 0.98 (0.57, 1.69) |
| SD units | **0.77 (0.70, 0.84)** | **0.80 (0.72, 0.87)** | **0.78 (0.68, 0.89)** | **0.81 (0.74, 0.90)** | **0.84 (0.76, 0.93)** | **0.84 (0.70, 0.99)** |
| HEART FAILURE |  |  |  |  |  |  |
| Social Networks |  |  |  |  |  |  |
| Low (referent) | 1.0 | 1.0 | 1.0 | 1.0 | 1.0 | 1.0 |
| High | **0.67 (0.48, 0.95)** | **0.61 (0.43, 0.86)** | 0.68 (0.40, 1.15) | 0.69 (0.42, 1.14) | 0.69 (0.42, 1.15) | 0.87 (0.36, 2.09) |
| SD units | **0.78 (0.67, 0.90)** | **0.77(0.67, 0.89)** | **0.78 (0.63, 0.95)** | 0.90 (0.75, 1.07) | 0.89 (0.74, 1.06) | 0.89 (0.67, 1.20) |

Model 1-age, education Model 2- Model 1+ physical activity, alcohol, smoking, dietary fat Model 3- Model 2+ hypertension, obesity, diabetes, depressive symptoms Abbreviations HR, hazard ratio; CI, confidence interval; SD, standard deviation **Boldface** = significant p <0.05

**Tables 4** and **5** show the results of mediation tests in the association of SN and CHD and SN and HF (stroke did not have a significant main effect), by sex. Natural indirect effects were not statistically significant for any of the mediators in the association of high (vs. low) SN and CHD or SN and HF for men or women. We additionally explored mediation of ideal smoking, ideal physical activity, and ideal nutrition and only found marginal indirect effects for ideal smoking for men and women **(Supplemental Tables A & B)**.

**Table 4.**
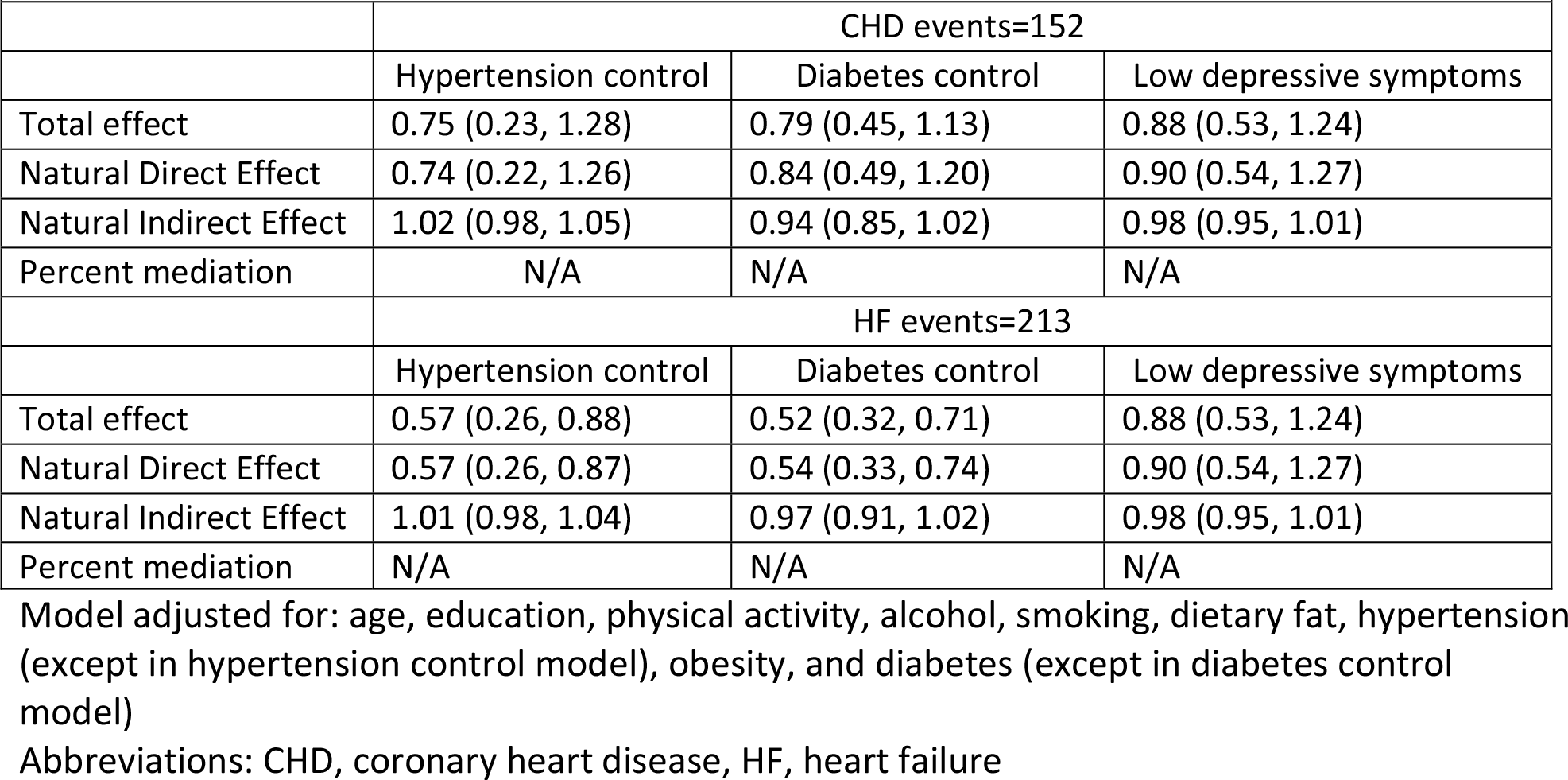
Effects from mediation tests of hypertension control, diabetes control and depressive symptoms in the association of high (vs. low) social network score and CHD and HF among women, Jackson Heart Study

**Table 5.** Effects from mediation tests of hypertension control, diabetes control and depressive symptoms in the association of high (vs. low) social network score and CHD and HF among men, Jackson Heart Study

|  |  |  |  |
| --- | --- | --- | --- |
|  | CHD events=110 |  |  |
|  | Hypertension control | Diabetes control | Low depressive symptoms |
| Total effect | 0.75 (0.23, 1.27) | 0.79 (0.45, 1.13) | 0.89 (0.53, 1.25) |
| Natural Direct Effect | 0.74 (0.22, 1.26) | 0.84 (0.49, 1.20) | 0.90 (0.54, 1.27) |
| Natural Indirect Effect | 1.01 (0.99, 1.03) | 0.94 (0.85, 1.02) | 0.98 (0.97, 1.01) |
| Percent mediation | N/A | N/A | N/A |
|  | HF events=118 |  |  |
|  | Hypertension control | Diabetes control | Low depressive symptoms |
| Total effect | 0.57 (0.26, 0.87) | 0.52 (0.32, 0.71) | 0.88 (0.53, 1.24) |
| Natural Direct Effect | 0.57 (0.26, 0.87) | 0.54 (0.33, 0.74) | 0.90 (0.54, 1.27) |
| Natural Indirect Effect | 1.01 (0.98, 1.04) | 0.97 (0.91, 1.02) | 0.98 (0.95, 1.01) |
| Percent mediation | N/A | N/A | N/A |

## Discussion

This study examined the association between SN and incident CHD, stroke, and HF among African American men and women enrolled in the JHS. In addition, we explored mediation of hypertension control, diabetes control, and depressive symptoms in the association between high (vs. low) SN and incident CHD, stroke, and HF. We found partial support for our hypotheses. There were stronger inverse associations in women vs. men for SN and incidence of CVD, specifically for CHD and HF; however, reporting better control of hypertension and diabetes and lower depressive symptomatology did not mediate the association of social networks and CHD or HF.

The inverse association between social connections and incident CVD, especially CHD, reported in this study is consistent with previous reports.^26, 27^ Our results highlight sex differences, with stronger associations noted for women than men. Rutledge et. al^8^ found a significant inverse association between SN and stroke among women from the Women’s Ischemia Syndrome Evaluation (WISE) study. Similar to our study, the association attenuated after adjustment for CVD risk factors. Among women from the WISE who had suspected coronary artery disease (CAD), Rutledge et al.^28^ also reported that women with higher SN scores showed a consistent pattern of reduced CAD. While there are few studies reporting associations between SN and HF events, studies have found reported associations between social isolation and HF among men and women. ^27, 29^

We explored whether having better control of risk factors mediated the association of SN and CHD and HF, but found no evidence of mediation. Studies have suggested that greater SN improves health behaviors, such as smoking and physical activity, which reduces the risk of CVD.^6, 30, 31^ Women from JHS were less likely to be alcoholic and were more likely to have ideal-smoking behaviors, but men were more likely to be physically active. While we explore smoking, physical activity and dietary habits as mediators, there were only marginal indirect effects observed. Perhaps SN offer other positive benefits, such as lower stress and greater tangible support, which leads to lower downstream CVD risk. Future work may need to evaluate psychosocial factors as potential mediators in the pathway between SN and CVD.

The results of this study should be interpreted with consideration of limitations. Generalizability is limited due to the analytic sample only including African American participants from Jackson, MS. There were reductions to the sample size due to loss to follow-up or death. However, inclusion of a large sample of African American participants, who were followed for over 10 years, in a study of social connectedness and CVD are strengths. Other strengths include use of surveillance data to capture CVD events and use of time-to-event analyses which considers temporality. In addition, use of formal mediation tests to understand pathways is another strength of this study.

A positive association between SN and CVD events were found more strongly among women. The results align with other studies conducted in women, which has implications for future public health interventions seeking to improve CVD burden and incidence in populations. Future work should continue to explore factors in the pathway of SN and a lower risk of CVD among African American adults, and the extent to which resilience measures, besides SN, may be protective of CVD risk in men. A formal CVD intervention targeting diet, morbidities, and physical activity paired with increasing social connectedness may be important for prevention of downstream CHD and HF in women.

## Data Availability

The data used for this study can be requested for purposes of reproducing results. Request to access this data set (or other data in the JHS) may be directed to the qualified researchers trained in human subject confidentiality within the JHS Coordinating Center at.

## Acknowledgements and Sources of funding

The Jackson Heart Study (JHS) is supported and conducted in collaboration with Jackson State University (HHSN268201800013I), Tougaloo College (HHSN268201800014I), the Mississippi State Department of Health (HHSN268201800015I) and the University of Mississippi Medical Center (HHSN268201800010I, HHSN268201800011I and HHSN268201800012I) contracts from the National Heart, Lung, and Blood Institute (NHLBI) and the National Institute for Minority Health and Health Disparities (NIMHD). The authors also wish to thank the staffs and participants of the JHS. Dr. Sims was also supported by the grant U54MD008176 from the NIMHD and 15SFDRN26140001 from the American Heart Association. Dr. Glover was supported by the Genetic Epidemiology of Heart, Lung, and Blood Traits Training Grant (GENHLB) T32 HL129982 and by the NHLBI under award F31HL159910.

## Disclaimer

The views expressed in this manuscript are those of the authors and do not represent the views of the National Heart, Lung, and Blood Institute; the National Institutes of Health; or the U.S. Department of Health and Human Services.

## Supplemental Figures

**Supplemental Figure A:**
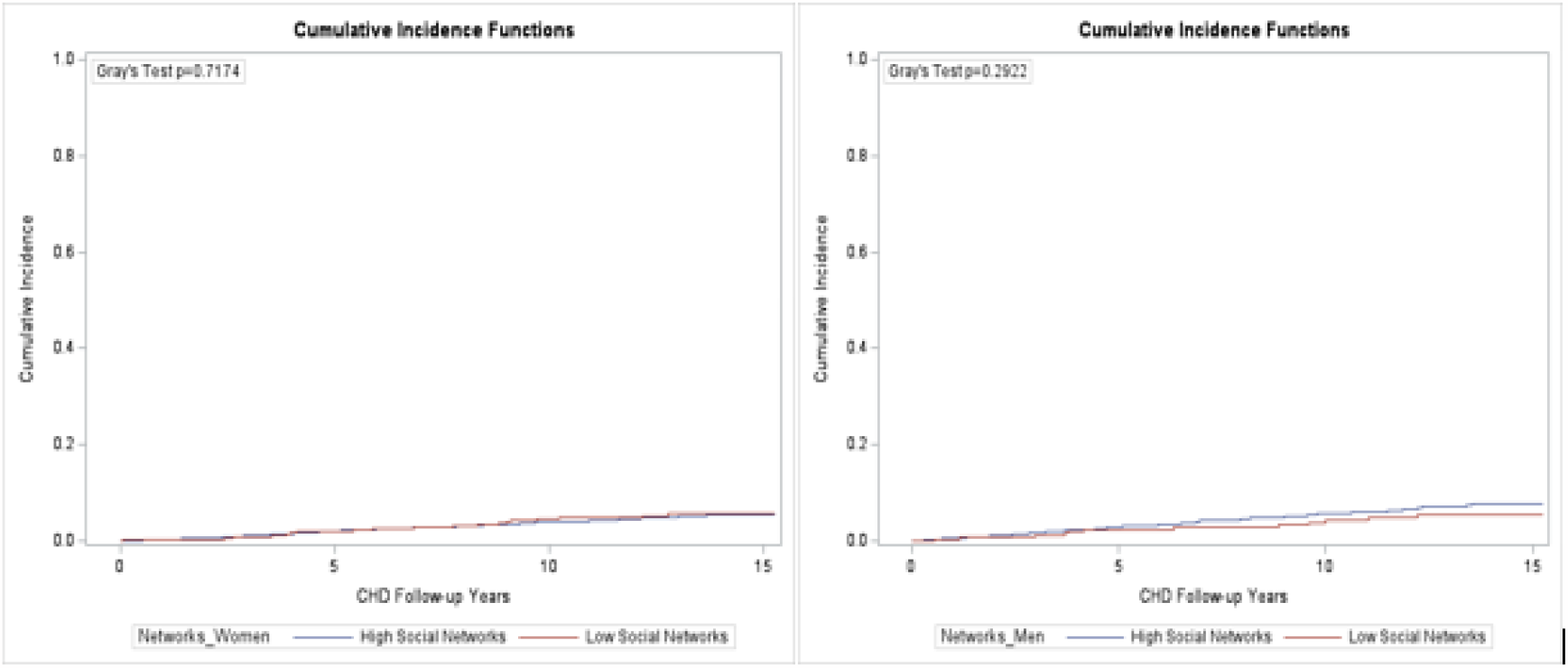
Kaplan-Meier Curve for CHD among women (left) and men (right).

**Supplemental Figure B:**
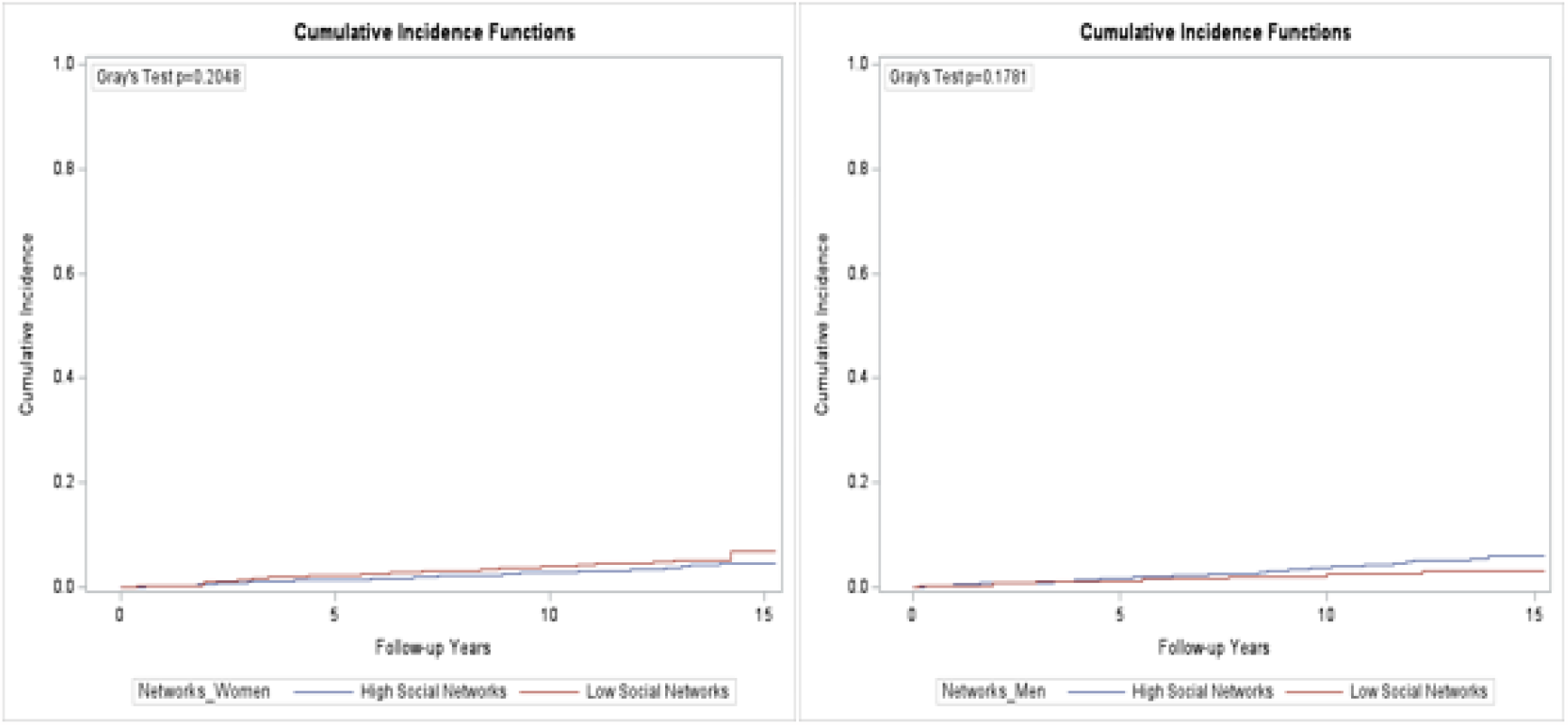
Kaplan-Meier Curve for stroke among women (left) and men (right).

## Supplemental tables

**Supplemental Table A.** Effects from mediation tests ideal smoking, ideal physical activity, and ideal nutrition in the association of high (vs. low) social network score and CHD and HF among women, Jackson Heart Study

|  |  |  |  |
| --- | --- | --- | --- |
|  | CHD events=110 |  |  |
|  | Ideal Smoking | Ideal Physical Activity | Ideal Nutrition |
| Total effect | 0.70 (0.41, 0.99) | 0.90 (0.54, 1.27) | 0.92 (0.54, 1.31) |
| Natural Direct Effect | 0.70 (0.41, 1.00) | 0.90 (0.54, 1.27) | 0.92 (0.54, 1.31) |
| Natural Indirect Effect | 0.98 (0.97, 1.00) | 1.00 (0.99, 1.00) | 1.00 (0.99, 1.01) |
| Percent mediation | N/A | N/A | N/A |
|  | HF events=118 |  |  |
|  | Ideal Smoking | Ideal Physical Activity | Ideal Nutrition |
| Total effect | 0.57 (0.35, 0.79) | 0.56 (0.36, 0.77) | 0.59 (0.37, 0.82) |
| Natural Direct Effect | 0.58 (0.36, 0.80) | 0.57 (0.36, 0.77) | 0.60 (0.40, 0.82) |
| Natural Indirect Effect | 0.99 (0.98, 1.00) | 1.00 (0.98, 1.02) | 1.00 (0.98, 1.01) |
| Percent mediation | N/A | N/A | N/A |

Note: Model adjusted for: age sex, education, physical activity (except in physical activity model), smoking (except in smoking model), alcohol, dietary fat, obesity, hypertension, diabetes, and depressive symptoms

**Supplemental Table B.** Effects from mediation tests ideal smoking, ideal physical activity, and ideal nutrition in the association of high (vs. low) social network score and CHD and HF among men, Jackson Heart Study

|  |  |  |  |
| --- | --- | --- | --- |
|  | CHD events=110 |  |  |
|  | Ideal Smoking | Ideal Physical Activity | Ideal Nutrition |
| Total effect | 0.72 (0.43, 1.01) | 0.90 (0.54, 1.27) | 0.92 (0.54, 1.31) |
| Natural Direct Effect | 0.74 (0.47, 1.04) | 0.90 (0.54, 1.27) | 0.92 (0.54, 1.31) |
| Natural Indirect Effect | 0.98 (0.96, 1.00) | 1.00 (0.99, 1.00) | 1.00 (0.99, 1.01) |
| Percent mediation | N/A | N/A | N/A |
|  | HF events=118 |  |  |
|  | Ideal Smoking | Ideal Physical Activity | Ideal Nutrition |
| Total effect | 0.57 (0.36, 0.78) | 0.56 (0.36, 0.77) | 0.59 (0.37, 0.82) |
| Natural Direct Effect | 0.57 (0.36, 0.79) | 0.57 (0.36, 0.77) | 0.59 (0.40, 0.79) |
| Natural Indirect Effect | 0.99 (0.97, 1.01) | 1.00 (0.97, 1.02) | 1.00 (0.98, 1.01) |
| Percent mediation | N/A | N/A | N/A |

Note: Model adjusted for: age sex, education, physical activity (except in physical activity model), smoking (except in smoking model), alcohol, dietary fat, obesity, hypertension, diabetes, and depressive symptoms

